# Patient Attitudes Toward Artificial Intelligence in Jordanian Healthcare: A Cross-Sectional Survey Study

**DOI:** 10.64898/2026.02.22.26346852

**Authors:** Zeena Al-Dabbas, Laith Khandakji, Nour Al-Shatarat, Hala Alqaisiah, Yazan Ibrahim, Tala Awed, Harith Baik, Mohammad Dawoud, Raghad Al-Haj Ali, Zaid Telfah, Yamamah Al-Hmaid, Adham Alsharkawi

## Abstract

Artificial intelligence (AI) is increasingly integrated into healthcare delivery, yet patient acceptance in resource constrained settings remains incompletely characterized. This study assessed attitudes toward AI supported care among patients attending hospitals in three Jordanian governorates (Amman, Balqa, Irbid) and examined demographic and digital literacy correlates of acceptance. In a cross sectional survey (n = 500 complete questionnaires), participants rated exposure to AI in healthcare and five attitudinal domains, namely perceived usefulness or performance expectancy, trust and transparency, privacy and perceived risks, empathy and human interaction, and readiness or behavioral intention, using 25 items on 5 point Likert scales. Patients expressed conditional optimism: empathy and human interaction was most strongly endorsed (M = 4.33, SD = 0.58), alongside relatively high perceived usefulness (M = 3.97, SD = 0.68), while trust and transparency (M = 3.57, SD = 0.74) and readiness (M = 3.66, SD = 0.90) were moderate to high; privacy and risk concerns were moderate (M = 3.51, SD = 0.77) and self reported exposure was lowest (M = 2.57, SD = 1.07). The highest agreement item indicated preference for AI to work alongside physicians rather than be relied on alone (M = 4.47, SD = 0.81). Trust and transparency and perceived usefulness were positively associated with readiness (r = 0.48 and r = 0.44, respectively; p *<*.001), while privacy and perceived risks were negatively correlated with trust and usefulness. In multivariable regression adjusting for gender, age group, education, prior AI health app or device use, and self rated digital skill, lower educational attainment (less than high school and high school) predicted reduced readiness, whereas higher digital skill predicted increased readiness (R^2^ = 0.101). These findings suggest that implementation strategies in Jordan should emphasize human involvement alongside AI, transparent communication and governance, and interventions that build digital confidence and reduce readiness gaps linked to education.

**Author summary:** AI is increasingly used in healthcare, for example to support diagnosis, triage, and treatment decisions. Whether these tools are accepted by patients depends not only on how well they work, but also on whether patients trust them, understand how they are used, and feel their privacy is protected. Evidence on patient views in middle income and resource constrained settings is still limited.

We surveyed 500 patients attending hospitals in three Jordanian governorates to understand how they view AI supported care. Patients generally expected AI to be useful, but they strongly preferred that clinicians remain actively involved and that AI supports rather than replaces physicians. Trust and perceived usefulness were closely linked to willingness to accept AI enabled care, while privacy concerns were present and shaped trust. Readiness to accept AI was lower among participants with lower educational attainment and higher among those with greater self rated digital skill. These findings suggest that successful implementation in Jordan should prioritize transparent communication, strong privacy safeguards, and human centered workflows, while also strengthening digital confidence to avoid widening gaps in acceptance.

## Introduction

AI in health care is increasingly being deployed as clinical tools that support decision-making and care workflows, with a prevailing view that AI should augment clinicians rather than replace professional judgment or accountability [1]. Advances in generative and multimodal foundation models have expanded capabilities while also broadening the risk surface, raising expectations for safety, transparency, privacy protection, and accountability across the system lifecycle [2, 3]. Regulators are clarifying lifecycle oversight for AI-enabled software functions that may change after authorization, including predetermined change control plans to support iterative improvement while maintaining reasonable assurance of safety and effectiveness [4].

Credible adoption depends on rigorous clinical evaluation and transparent reporting. DECIDE-AI provides guidance for early-stage clinical studies of AI decision-support systems [5], and newer reporting standards extend requirements to clinical prediction models, diagnostic accuracy studies using AI, and studies involving large language models [6–8]. Because these systems influence patient-related decisions, safe use requires appropriate trust calibration and reliance [9]. A systematic review indicates that patients and the public often anticipate benefits from clinical AI but prefer human oversight and report concerns about transparency and accountability [10]. Evidence of proxy-driven bias in widely used health algorithms shows that equity risks are practical rather than hypothetical, motivating context-specific assessment of benefit, trust, privacy expectations, and effects on the clinician-patient relationship [11].

### Recent literature on attitudes toward AI-supported care

Across settings, the literature converges on a consistent conclusion: stakeholders often view AI in healthcare as promising, but acceptance is typically conditional, shaped by perceived usefulness, trust and transparency, privacy and perceived risks, empathy and human interaction, and readiness or behavioral intention [12, 13]. Framing prior evidence around these constructs provides a direct rationale for the attitudinal domains assessed in the present study within Jordan.

Perceived clinical value, especially improved accuracy, efficiency, or timeliness, is a recurrent driver of acceptance. In a large multinational survey of hospital patients across diverse health systems, respondents generally supported AI use but expressed preferences indicating that AI should add value without displacing clinician judgment [14]. Experimental evidence similarly suggests that acceptance increases when AI is presented as more accurate and when recommendations are communicated in ways that emphasize patient centered benefit [15]. In the United States, synthesis of nationally representative surveys indicates that many members of the public expect medicine to benefit from AI, while support varies across applications and population subgroups [13]. At the same time, expectations regarding whether AI will improve affordability and access may be modest and closely tied to broader trust in the healthcare system [16]. Collectively, these findings imply that perceived usefulness is not a generic enthusiasm for technology; it depends on whether AI is understood as improving outcomes patients care about, such as diagnostic reliability, access, and convenience, and whether those benefits are communicated credibly.

Trust emerges as both a direct determinant of acceptance and a mechanism through which other concerns are filtered. Multiple patient studies show that individuals are more comfortable with AI when it is positioned as clinician supervised support rather than autonomous decision making [14, 17]. Qualitative work in wound care reinforces that limited AI understanding is common, and that trust often operates through clinician mediation: patients prefer clinicians to interpret and contextualize AI outputs, while transparency and alignment with professional judgment strengthen confidence [18]. From the clinician side, surveys emphasize governance concerns, including accountability, confidentiality, and bias, and support the norm that patients should retain access to human clinical judgment and second opinions [19]. These patterns motivate measuring transparency and trust together; in practice, trust frequently depends on whether AI use is explainable, supervised, and embedded within established clinical responsibility.

Even when general attitudes are positive, privacy and risk concerns remain among the most durable barriers. Public evidence in the United States shows that comfort with AI does not necessarily translate into comfort with data access, and that perceived bias and discrimination risks may meaningfully shape acceptance for some groups [13, 20]. Clinician perspectives similarly underscore confidentiality and bias as persistent concerns that require clear regulatory and ethical frameworks [19]. These concerns are directly relevant to measuring perceived risks and privacy attitudes among patients, particularly in contexts where public discussion of data governance and accountability may be limited.

A distinct and repeatedly observed constraint on acceptance is fear of reduced interpersonal care. In sensitive treatment contexts, such as radiotherapy, patients may view AI implementation more negatively when it is perceived to diminish personal interaction or relational support [21]. In the Arab region, student and trainee samples also express concerns that AI lacks empathy and may not adapt flexibly to individualized patient needs [22]. This construct is especially important for patient facing services in which reassurance, communication, and dignity are central to perceived quality of care.

Across stakeholders, enthusiasm often coexists with limited knowledge and limited formal preparation. A systematic review and meta analysis across medical, dental, and nursing student populations found generally positive attitudes alongside only moderate knowledge, implying an enthusiasm preparedness gap that may influence how confidently future clinicians communicate about AI to patients [23]. Similar patterns are reported in specific trainee groups, including radiology residents [24] and nursing students [25]. Because patient acceptance is partly shaped by clinical communication and implementation quality, readiness within the healthcare workforce provides critical context even in patient centered studies.

Arab region studies broadly align with international findings but highlight two recurring themes: strong optimism about AI potential and substantial variability in knowledge and comfort. Medical student surveys in Kuwait and multi country Arab samples report high perceived importance of AI and strong support for AI education, yet also document pronounced knowledge deficits [26, 27]. In Lebanon, similar gaps between awareness and deeper understanding have been reported [28]. Patient and public facing data in the region remain comparatively limited and often concentrate on radiology or specific implementations. In Saudi Arabia, patients expressed interest in AI supported radiology but still valued direct interaction with radiologists [29], while public samples demonstrate generally positive orientations tempered by concerns about safety, trust, regulation, and workforce impact [30]. Evidence from the United Arab Emirates also indicates that public perceptions can be mixed and may skew unfavorable in some samples, reinforcing the need for context sensitive measurement rather than assuming uniformly positive attitudes [31]. These regional patterns support measuring both perceived usefulness and relational and ethical concerns, including privacy, empathy, and transparency, when assessing acceptance.

Jordan specific evidence to date has largely emphasized clinicians and students rather than patients, consistently reporting interest in AI alongside barriers related to training, resources, and governance. A survey of healthcare professionals in Jordan documented perceptions of AI alongside perceived barriers and risks [32]. Nursing focused work similarly links AI related knowledge, attitudes, practices, and perceived barriers to workforce outcomes such as intent to stay [33]. Among medical and allied health students, studies commonly report moderate knowledge, strong interest, and calls for curricular integration of AI [34–37]. Specialty focused samples, including radiology and anesthesiology, further illustrate cautious optimism coupled with concerns about role change and human interaction [38, 39].

Despite this growing stakeholder literature, direct evidence on patients’ attitudes within Jordan, and critically, how key constructs such as usefulness, trust and transparency, privacy and perceived risks, and empathy and human interaction relate to behavioral intention across demographic and digital literacy groups, remains limited. This gap motivates the present study’s focus on patient attitudes within the Jordanian healthcare system and their relationship to acceptance of AI supported care.

## Materials and Methods

### Study design and setting

We conducted a cross-sectional questionnaire survey among a convenience sample of 500 adults recruited from seven tertiary public and academic teaching hospitals across three Jordanian governorates (Amman, Balqa, and Irbid). Data collection took place between July and October 2025.

### Participants and data collection

Eligible participants included adults (aged ≥18 years) attending the participating hospitals who were capable of providing informed consent. The study was approved by the Institutional Review Board (IRB) of Al-Balqa Applied University (Approval No. 2026/2025/2/25). Recruitment was conducted in outpatient clinics, inpatient wards, and waiting areas using a convenience sampling approach. Trained research staff approached potential participants to explain the study’s purpose and obtain informed consent, which was documented by checking a box on the electronic form. The questionnaire was administered in Arabic by the research staff using electronic devices (smartphones or tablets).

Prior to the main study, the questionnaire was pilot tested with 44 participants from the participating hospitals. Feedback from the pilot test and internal consistency analysis were used to refine item wording and improve clarity before final data collection.

Demographic and background variables included age group, gender, highest education level, governorate, hospital (selected from a standardized drop-down list), prior use of AI-based health applications or devices (Yes/No/Not sure), and self-rated digital skill (Low/Moderate/High).

### Measures

Attitudes toward AI in healthcare were measured using 25 Likert-type items rated on a 5-point scale (1 = strongly disagree, 2 = disagree, 3 = not sure, 4 = agree, 5 = strongly agree). Items were grouped a priori into six constructs:

- Exposure to AI in healthcare (2 items)
- Trust & transparency (5 items)
- Perceived usefulness / performance expectancy (5 items)
- Privacy & perceived risks (5 items)
- Empathy & human interaction (5 items)
- Readiness / behavioral intention (3 items)

Composite scores were calculated as the mean of items within each construct. Higher scores indicated greater exposure, higher trust, higher perceived usefulness, greater privacy/risk concern, stronger preference for empathy/human interaction, and higher readiness/behavioral intention, respectively.

### Statistical analysis

Data integrity was assessed by checking value ranges, internal consistency of coding, and completeness of responses. There were no missing item responses, so all analyses were conducted on complete cases. Internal consistency of each construct was evaluated using Cronbach’s alpha (*α*).

For descriptive statistics, we calculated means (M), standard deviations (SD), and 95% confidence intervals for each item and each construct. Group differences in construct scores were examined using Welch’s *t*-tests for two-group comparisons (e.g., gender, prior AI health app/device use [Yes vs No, excluding “Not sure”]) and one-way analysis of variance (ANOVA) for age group, education level, and self-rated digital skill (Low/Moderate/High), with Tukey post-hoc comparisons where appropriate. Effect sizes were reported as Hedges’ *g* for *t*-tests and *η*^2^ for ANOVA.

Pearson correlation coefficients were computed to quantify associations among the construct scores (for example, trust, perceived usefulness, privacy or risk, empathy or human interaction, exposure, and readiness or behavioral intention). To examine independent associations with behavioral intention, we fitted a multiple linear regression model with the Readiness or behavioral intention composite as the dependent variable and gender, age group, education level, prior AI health application or device use, and self rated digital skill (coded 1 = Low, 2 = Moderate, 3 = High) as predictors. Statistical significance was defined as *p* < 0.05 using two sided tests. Given the exploratory aim of the study, no formal adjustment was made for multiple comparisons. Analyses were conducted in Python (version 3.12) using the pandas, SciPy, and statsmodels libraries.

## Results

### Data integrity and preparation

The final dataset comprised 500 complete questionnaires with no missing values across 34 variables (9 demographic/administrative fields and 25 Likert-scale attitude items). All Likert-type responses fell within the prespecified range of 1–5, and no out-of-range or implausible values were identified.

### Sample characteristics

Participant characteristics are summarized in Table 1. The sample was 51.6% female (n=258). The largest age group was < 25 years (26.8%), and the most common highest education level was a Bachelor’s degree (45.4%). Most participants were recruited from hospitals in Amman (45.6%), followed by Balqa (31.8%) and Irbid (22.6%). Just under half of respondents reported no prior use of AI-based health applications/devices (47.8%), and the majority rated their digital skill as moderate (60.2%).

**Table 1.** Participant demographics (N=500).

| Variable | Category | n | % |
| --- | --- | --- | --- |
| Age group | < 25 | 134 | 26.8 |
| Age group | 25–34 | 105 | 21.0 |
| Age group | 35–44 | 95 | 19.0 |
| Age group | 45–54 | 74 | 14.8 |
| Age group | 55–64 | 68 | 13.6 |
| Age group | ≥ 65 | 24 | 4.8 |
| Gender | Female | 258 | 51.6 |
| Gender | Male | 242 | 48.4 |
| Education | Bachelor's | 227 | 45.4 |
| Education | High school | 118 | 23.6 |
| Education | Diploma/College | 75 | 15.0 |
| Education | <High school | 32 | 6.4 |
| Education | Master's | 25 | 5.0 |
| Education | Doctorate | 23 | 4.6 |
| Governorate | Amman | 228 | 45.6 |
| Governorate | Balqa | 159 | 31.8 |
| Governorate | Irbid | 113 | 22.6 |
| Hospital | Al-Hussein Al-Salt New Hospital | 192 | 38.4 |
| Hospital | Jordan University Hospital | 124 | 24.8 |
| Hospital | King Abdullah University Hospital | 65 | 13.0 |
| Hospital | Princess Basma Teaching Hospital | 50 | 10.0 |
| Hospital | Prince Hamzah Hospital | 50 | 10.0 |
| Hospital | Al-Hussein Medical City | 17 | 3.4 |
| Hospital | Al-Salt Hospital | 2 | 0.4 |
| Prior AI health app/device use | No | 239 | 47.8 |
| Prior AI health app/device use | Yes | 215 | 43.0 |
| Prior AI health app/device use | Not sure | 46 | 9.2 |
| Self-rated digital skill | Moderate | 301 | 60.2 |
| Self-rated digital skill | High | 136 | 27.2 |
| Self-rated digital skill | Low | 63 | 12.6 |

### Reliability (internal consistency)

Internal consistency estimates for the six attitude constructs are shown in Table 2. Trust & transparency (*α* = 0.745) and Perceived usefulness / performance expectancy (*α* = 0.756) demonstrated acceptable internal consistency, indicating reasonably coherent measurement of these domains. The remaining constructs showed more modest reliability (Exposure *α* = 0.596; Privacy & perceived risks *α* = 0.665; Empathy & human interaction *α* = 0.637; Readiness / behavioral intention *α* = 0.621), suggesting greater heterogeneity in responses and that these composite scores should be interpreted with appropriate caution.

**Table 2.** Internal consistency reliability (Cronbach’s alpha) by construct.

| Construct | Items (*k*) | Cronbach's $\alpha$ |
| --- | --- | --- |
| Exposure to AI in healthcare | 2 | 0.596 |
| Trust & transparency | 5 | 0.745 |
| Perceived usefulness / performance expectancy | 5 | 0.756 |
| Privacy & perceived risks | 5 | 0.665 |
| Empathy & human interaction | 5 | 0.637 |
| Readiness / behavioral intention | 3 | 0.621 |

### Construct-level descriptive statistics

Construct-level descriptive statistics are presented in Table 3 and illustrated in Figure 1. Empathy & human interaction had the highest mean score (M=4.33, SD=0.58), indicating strong endorsement of the importance of human involvement in care even when AI is used. Perceived usefulness / performance expectancy was also relatively high (M=3.97, SD=0.68). Trust & transparency (M=3.57, SD=0.74) and Readiness / behavioral intention (M=3.66, SD=0.90) fell in the moderate-to-high range. Privacy & perceived risks had a moderate mean (M=3.51, SD=0.77), reflecting salient but not extreme concern about data security and potential harm. Exposure to AI in healthcare had the lowest mean (M=2.57, SD=1.07), indicating comparatively limited self-reported awareness and direct experience with AI-enabled services.

**Table 3.** Construct-level descriptive statistics (1–5 Likert).

| Construct | M | SD | 95% CI LL | 95% CI UL |
| --- | --- | --- | --- | --- |
| Exposure to AI in healthcare | 2.57 | 1.07 | 2.48 | 2.66 |
| Trust & transparency | 3.57 | 0.74 | 3.51 | 3.64 |
| Perceived usefulness / performance expectancy | 3.97 | 0.68 | 3.92 | 4.03 |
| Privacy & perceived risks | 3.51 | 0.77 | 3.44 | 3.58 |
| Empathy & human interaction | 4.33 | 0.58 | 4.28 | 4.38 |
| Readiness / behavioral intention | 3.66 | 0.90 | 3.58 | 3.73 |

**Fig 1.**
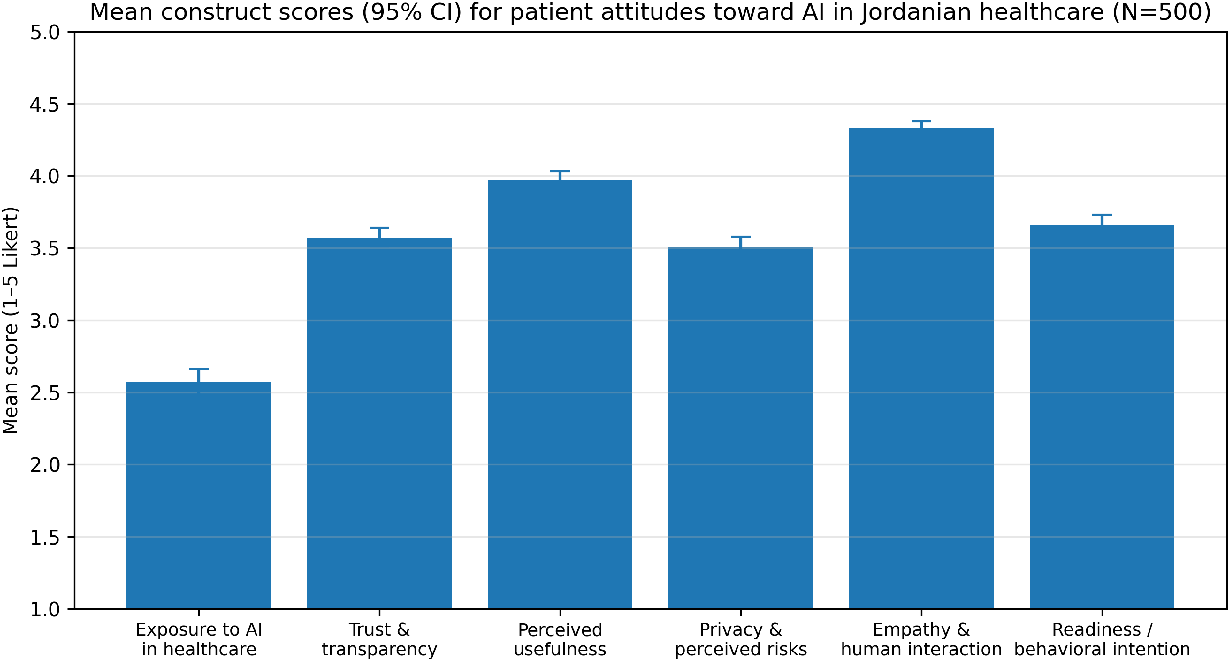
Mean construct scores (with 95% confidence intervals) for patient attitudes toward AI in Jordanian healthcare. Scores range from 1 (strongly disagree) to 5 (strongly agree).

### Item-level descriptive statistics

Item-level descriptive statistics are provided in Table 4. The item with the highest agreement was, “I prefer AI to work alongside the physician rather than relying on it alone” (Empathy/Human interaction; M=4.47, SD=0.81), indicating strong support for AI in a human-in-the-loop configuration. High agreement was also observed for items emphasizing the ongoing importance of human empathy and communication (e.g., preference for human explanation of test results). The lowest agreement was observed for, “Previously received AI-assisted examination/care” (Exposure; M=2.29, SD=1.24), consistent with limited self-reported prior exposure to AI-supported clinical services.

**Table 4.** Item-level descriptive statistics ranked from highest to lowest agreement. Construct codes: EXP=Exposure; TR=Trust/Transparency; PU=Perceived usefulness; PR=Privacy/Risk; HI=Human interaction; BI=Behavioral intention.

| Item | Construct | Item (English) | M | SD |
| --- | --- | --- | --- | --- |
| HI5 | HI | I prefer AI to work alongside the physician rather than relying on it alone. | 4.47 | 0.81 |
| HI1 | HI | Human empathy remains important to me even with AI-based diagnosis. | 4.39 | 0.89 |
| HI4 | HI | I feel reassured when a human explains my test results rather than a device. | 4.38 | 0.90 |
| HI3 | HI | Heavy reliance on AI may reduce human connection between doctors and patients. | 4.25 | 0.93 |
| BI2 | BI | I prefer a shared approach where AI and the physician jointly review my case. | 4.22 | 0.85 |
| PU2 | PU | AI can speed up delivery of medical care for patients like me. | 4.17 | 0.84 |
| HI2 | HI | AI cannot understand the emotional aspects of health-care. | 4.17 | 1.02 |
| PU1 | PU | AI can help physicians detect diseases early. | 4.11 | 0.87 |

| Item | Construct | Item (English) | M | SD |
| --- | --- | --- | --- | --- |
| PU5 | PU | AI diagnosis would allow physicians more time for complex tasks. | 4.03 | 0.93 |
| PU4 | PU | AI can improve the quality of care I receive. | 3.85 | 0.98 |
| TR5 | TR | Would feel comfortable relying on AI if recommended by my physician. | 3.85 | 0.98 |
| PR3 | PR | Errors in AI diagnosis could lead to serious health risks for patients. | 3.81 | 1.00 |
| PR5 | PR | Concerned AI recommendations could override the treating physician's judgment. | 3.75 | 1.03 |
| TR3 | TR | Trust AI recommendations more when AI/doctor explains reasons for them. | 3.72 | 1.08 |
| PU3 | PU | Using AI contributes to lowering treatment costs. | 3.70 | 1.05 |
| TR1 | TR | Trust healthcare providers will use AI responsibly. | 3.65 | 1.03 |
| TR4 | TR | Trust AI systems in this hospital will not cause me harm. | 3.49 | 1.07 |
| PR4 | PR | Difficult to correct mistakes if AI makes a wrong decision about my health. | 3.47 | 1.10 |
| BI3 | BI | If AI proven more accurate than doctors, I would accept an AI diagnosis. | 3.41 | 1.35 |
| TR2 | TR | Believe AI systems used here are accurate in diagnosing diseases. | 3.35 | 1.06 |
| BI1 | BI | If AI as accurate as doctor, I would accept it conducting initial assessment. | 3.34 | 1.30 |
| PR2 | PR | Fear personal data may be shared without consent through AI systems. | 3.33 | 1.30 |
| PR1 | PR | Concerned my medical data may not be secure if used by AI. | 3.18 | 1.31 |
| EXP2 | EXP | Aware that AI is currently used in some hospital departments. | 2.85 | 1.30 |
| EXP1 | EXP | Previously received AI-assisted examination/care. | 2.29 | 1.24 |

### Group differences in composite scores

#### Gender

Gender differences in construct scores are summarized in Table 5. Welch’s *t*-tests indicated that females reported higher exposure/awareness than males (Exposure: *t*(485.4) = 2.16, *p* =.031, Hedges’ *g* = 0.19), reflecting a small difference in favor of women. In contrast, males reported higher perceived usefulness than females (Usefulness: *t*(486.7) = −2.23, *p* =.027, Hedges’ *g* = −0.20), also a small effect. No statistically significant gender differences were observed for trust, privacy/risk, empathy/human interaction, or readiness/behavioral intention (all *p* >.05).

**Table 5.** Gender differences (Welch’s *t*-tests) on composite construct scores.

| Composite score | Female M (SD) | Male M (SD) | *t*(df) | *p* | Hedges' *g* |
| --- | --- | --- | --- | --- | --- |
| Exposure to AI in healthcare | 2.67 (1.02) | 2.46 (1.12) | 2.16 (485.4) | .031 | 0.19 |
| Trust & transparency | 3.54 (0.71) | 3.61 (0.78) | -1.15 (486.5) | .250 | -0.10 |
| Usefulness / performance expectancy | 3.91 (0.64) | 4.04 (0.70) | -2.23 (486.7) | .027 | -0.20 |
| Privacy & perceived risks | 3.52 (0.72) | 3.50 (0.82) | 0.39 (480.0) | .700 | 0.03 |
| Empathy & human interaction | 4.30 (0.60) | 4.37 (0.56) | -1.27 (497.9) | .205 | -0.11 |
| Readiness / behavioral intention | 3.59 (0.91) | 3.73 (0.88) | -1.75 (497.3) | .082 | -0.16 |

#### Age group

Age-group differences are shown in Table 6. One-way ANOVA indicated statistically significant variation across age groups for Exposure (*F* (5, 494) = 2.62, *p* =.024, *η*^2^ = 0.026), Trust & transparency (*F* (5, 494) = 3.10, *p* =.009, *η*^2^ = 0.030), and Empathy & human interaction (*F* (5, 494) = 2.35, *p* =.040, *η*^2^ = 0.023), with small effect sizes. Tukey post-hoc comparisons showed that participants aged ≥65 reported lower exposure and lower trust than those aged < 25 years (*p* =.035 and *p* =.022, respectively). Participants aged 55–64 reported a higher preference for human interaction than those aged 35–44 (*p* =.045). No significant age-group differences were detected for perceived usefulness, privacy/risk, or readiness/behavioral intention (all *p* >.05).

**Table 6.**
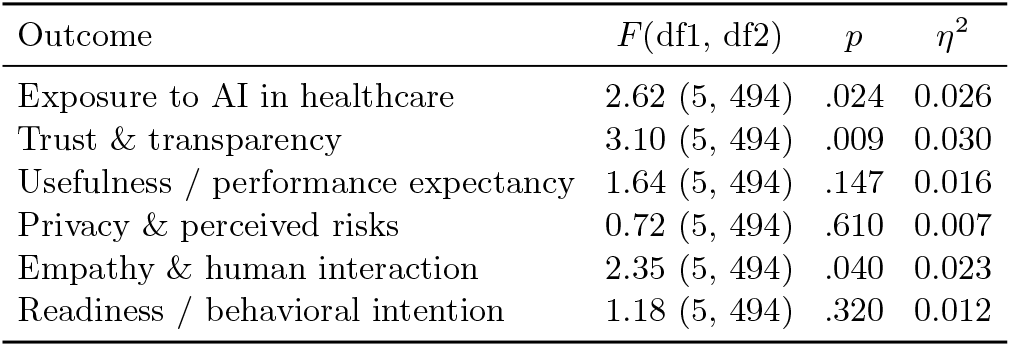
Age-group differences in composite scores (one-way ANOVA).

| Outcome | $F(df1, df2)$ | $p$ | $\eta^2$ |
| --- | --- | --- | --- |
| Exposure to AI in healthcare | 2.62 (5, 494) | .024 | 0.026 |
| Trust & transparency | 3.10 (5, 494) | .009 | 0.030 |
| Usefulness / performance expectancy | 1.64 (5, 494) | .147 | 0.016 |
| Privacy & perceived risks | 0.72 (5, 494) | .610 | 0.007 |
| Empathy & human interaction | 2.35 (5, 494) | .040 | 0.023 |
| Readiness / behavioral intention | 1.18 (5, 494) | .320 | 0.012 |

#### Education

Associations between education level and construct scores are summarized in Table 7. Education was significantly associated with Exposure (*F* (5, 494) = 9.38, *p* <.001, *η*^2^ = 0.087), Trust (*F* (5, 494) = 2.44, *p* =.033, *η*^2^ = 0.024), Perceived usefulness (*F* (5, 494) = 2.86, *p* =.015, *η*^2^ = 0.028), Privacy/Risk (*F* (5, 494) = 2.55, *p* =.027, *η*^2^ = 0.025), and Readiness/behavioral intention (*F* (5, 494) = 6.88, *p* <.001, *η*^2^ = 0.065). Overall, participants with lower educational attainment tended to report lower exposure to AI in healthcare and lower readiness to accept AI-enabled care than those with a Bachelor’s degree. For Privacy/Risk, although the omnibus ANOVA was statistically significant, no pairwise comparisons remained significant after Tukey adjustment, suggesting that differences in privacy/risk perceptions between education groups were small.

**Table 7.** Education differences in composite scores (one-way ANOVA).

| Outcome | $F(df1, df2)$ | $p$ | $\eta^2$ |
| --- | --- | --- | --- |
| Exposure to AI in healthcare | 9.38 (5, 494) | < .001 | 0.087 |
| Trust & transparency | 2.44 (5, 494) | .033 | 0.024 |
| Usefulness / performance expectancy | 2.86 (5, 494) | .015 | 0.028 |
| Privacy & perceived risks | 2.55 (5, 494) | .027 | 0.025 |
| Empathy & human interaction | 1.32 (5, 494) | .254 | 0.013 |
| Readiness / behavioral intention | 6.88 (5, 494) | < .001 | 0.065 |

### Differences by prior AI use and self-rated digital skill

Differences in construct scores by prior AI health app/device use are presented in Table 8. Compared with participants who reported no prior use, those reporting prior use (excluding the “Not sure” category) had higher scores for Exposure (M=2.77 vs 2.35; *p* <.001; Cohen’s *d* = 0.40), Trust & transparency (M=3.75 vs 3.47; *p* <.001; *d* = 0.38), Perceived usefulness (M=4.10 vs 3.90; *p* =.002; *d* = 0.29), and Readiness / behavioral intention (M=3.80 vs 3.53; *p* =.002; *d* = 0.30). Effect sizes were small to small–moderate.

**Table 8.** Differences in composite scores by prior AI health app/device use (Yes vs No; excluding “Not sure”).

| Construct | No (n=239) M | Yes (n=215) M | $p$ | Cohen's $d$ |
| --- | --- | --- | --- | --- |
| Exposure | 2.35 | 2.77 | < .001 | 0.40 |
| Trust & transparency | 3.47 | 3.75 | < .001 | 0.38 |
| Usefulness / performance expectancy | 3.90 | 4.10 | .002 | 0.29 |
| Readiness / behavioral intention | 3.53 | 3.80 | .002 | 0.30 |

Self-rated digital skill showed graded differences across several constructs. For example, readiness to accept AI-enabled care differed significantly by digital skill (Low, Moderate, High: *F* (2, 497) = 15.11, *p* <.001, *η*^2^ = 0.057), with mean readiness scores increasing from Low (M=3.20) to Moderate (M=3.63) to High (M=3.92). Similar patterns were observed for exposure, trust, and perceived usefulness, whereas privacy/risk concerns and empathy/human interaction scores showed little systematic variation by digital skill or prior AI use.

### Inter-construct correlations

Inter-scale correlations among the six composite constructs are reported in Table 10. Trust & transparency and Perceived usefulness were moderately and positively correlated (Trust–Usefulness: *r* = 0.55, 95% CI [0.48, 0.61], *p* <.001). Both constructs were also positively correlated with Readiness / behavioral intention (Trust–Readiness: *r* = 0.48, 95% CI [0.41, 0.55], *p* <.001; Usefulness–Readiness: *r* = 0.44, 95% CI [0.37, 0.51], *p* <.001). Privacy & perceived risks was negatively correlated with Trust (Trust–Privacy/Risk: *r* = −0.23, 95% CI [−0.31, −0.15], *p* <.001) and with Perceived usefulness (Usefulness–Privacy/Risk: *r* = −0.17, *p* <.001). Privacy/Risk was positively associated with Empathy & human interaction (Privacy/Risk–Empathy/Human interaction: *r* = 0.18, 95% CI [0.10, 0.27], *p* <.001). Exposure to AI in healthcare was positively correlated with Trust (Exposure–Trust: *r* = 0.25, 95% CI [0.17, 0.33], *p* <.001) and with Readiness (Exposure–Readiness: *r* = 0.24, 95% CI [0.16, 0.32], *p* <.001). All reported correlations were statistically significant at *p* <.001.

### Regression predictors of behavioral intention

A multiple linear regression model was fitted with the Readiness / behavioral intention composite as the dependent variable and gender, age group, education level, prior AI health app/device use, and self-rated digital skill as predictors (Table 9). The overall model was statistically significant (*F* (14, 485) = 3.89, *p* <.001) and accounted for approximately 10.1% of the variance in readiness (*R*^2^ = 0.101).

**Table 9.** Multiple linear regression predicting behavioral intention (Readiness/Intention composite). Model: *F* (14, 485) = 3.89, *p* <.001, *R*^2^ =.101.

| Predictor | B | SE | $t$ | $p$ | 95% CI |
| --- | --- | --- | --- | --- | --- |
| Intercept | 3.245 | 0.199 | 16.30 | < .001 | [2.854, 3.637] |
| Male (vs Female) | 0.113 | 0.080 | 1.42 | .158 | [-0.044, 0.270] |
| Age < 25 (vs 25–34) | -0.025 | 0.115 | -0.22 | .825 | [-0.251, 0.201] |
| Age 35–44 (vs 25–34) | -0.110 | 0.125 | -0.88 | .379 | [-0.355, 0.135] |
| Age 45–54 (vs 25–34) | -0.014 | 0.137 | -0.10 | .920 | [-0.284, 0.256] |
| Age 55–64 (vs 25–34) | 0.005 | 0.141 | 0.04 | .971 | [-0.272, 0.282] |
| Age ≥ 65 (vs 25–34) | -0.083 | 0.205 | -0.40 | .686 | [-0.485, 0.319] |
| Education <High school (vs Bachelor's) | -0.512 | 0.171 | -2.99 | .003 | [-0.849, -0.175] |
| Education High school (vs Bachelor's) | -0.254 | 0.103 | -2.47 | .014 | [-0.456, -0.052] |
| Education Diploma/College (vs Bachelor's) | -0.189 | 0.122 | -1.56 | .120 | [-0.428, 0.050] |
| Education Master's (vs Bachelor's) | -0.198 | 0.195 | -1.02 | .309 | [-0.581, 0.184] |
| Education Doctorate (vs Bachelor's) | 0.326 | 0.203 | 1.60 | .110 | [-0.074, 0.725] |
| Prior AI use: Yes (vs No) | 0.105 | 0.089 | 1.17 | .241 | [-0.071, 0.280] |
| Prior AI use: Not sure (vs No) | -0.068 | 0.143 | -0.48 | .634 | [-0.348, 0.212] |
| Digital skill (1=Low to 3=High) | 0.217 | 0.072 | 3.02 | .003 | [0.076, 0.358] |

Compared with participants holding a Bachelor’s degree, those with <High school education (B= −0.512, SE=0.171, *p* =.003) and those with High school education (B= −0.254, SE=0.103, *p* =.014) had significantly lower readiness scores, adjusting for other covariates. Self-rated digital skill was positively associated with readiness (B=0.217 per one-unit increase on the 1–3 scale, SE=0.072, *p* =.003), indicating higher readiness among participants with greater digital confidence. In contrast, gender, age group, and prior AI health app/device use were not statistically significant predictors in the multivariable model (all *p* >.05).

## Discussion

In this Jordanian hospital-based sample, patients expressed conditional optimism toward the use of AI in healthcare. Perceived usefulness of AI was relatively high (M=3.97), and both trust in AI and associated systems (M=3.57) and readiness to accept AI-enabled care (M=3.66) were in the moderate-to-high range. At the same time, participants placed the strongest emphasis on empathy and human interaction (M=4.33), indicating that human involvement remains central to how care should be delivered (Table 3). Privacy and perceived-risk concerns were also salient (M=3.51), and self-reported exposure to AI in healthcare was comparatively low (M=2.57), consistent with limited direct experience. The highest-scoring item, a preference for AI to work alongside the physician rather than being relied on alone (M=4.47), underscores that acceptability was greatest when AI was framed as clinician-supportive rather than autonomous (Table 4).

Beyond mean levels, the pattern of associations between constructs helps clarify how acceptance is structured. Trust & transparency and Perceived usefulness were moderately and positively correlated, and both were positively associated with Readiness / behavioral intention (Table 10). These associations suggest that, within this sample, willingness to engage with AI-enabled care is closely linked to expectations of benefit and confidence in how AI will be used, rather than being a diffuse or purely attitudinal stance. Privacy & perceived risks were inversely associated with both trust and perceived usefulness, indicating that concerns about harm and data security are central to how patients evaluate AI. At the same time, privacy and risk concerns were only moderate on average, and the combination of moderate concern with moderate or high readiness suggests that worries about privacy do not necessarily prevent willingness to use AI when patients also perceive clear benefits and trust the systems and clinicians involved.

**Table 10.** Inter-scale correlations among composite construct scores.

| Pair | $r$ | 95% CI | $p$ |
| --- | --- | --- | --- |
| Trust – Usefulness | 0.55 | [0.48, 0.61] | < .001 |
| Trust – Readiness | 0.48 | [0.41, 0.55] | < .001 |
| Usefulness – Readiness | 0.44 | [0.37, 0.51] | < .001 |
| Trust – Privacy/Risk | -0.23 | [-0.31, -0.15] | < .001 |
| Usefulness – Privacy/Risk | -0.17 | — | < .001 |
| Privacy/Risk – Empathy/Human interaction | 0.18 | [0.10, 0.27] | < .001 |
| Exposure – Trust | 0.25 | [0.17, 0.33] | < .001 |
| Exposure – Readiness | 0.24 | [0.16, 0.32] | < .001 |

The findings also highlight factors that may be more amenable to intervention than demographic characteristics alone. Age- and gender-related differences in construct scores were statistically detectable in some cases but consistently small in magnitude (Tables 5 and 6), which limits the utility of demographic profiling as the primary lens for implementation. In contrast, educational attainment and digital skill showed more consistent associations with readiness. Participants with lower education reported lower readiness than those with a Bachelor’s degree, and in the multivariable model, both < High school and High school education remained independently associated with reduced readiness after adjustment for other covariates (Table 9). Self-rated digital skill showed a graded relationship with readiness and related constructs: participants who rated their digital skill as high reported higher exposure, trust, perceived usefulness, and readiness than those with lower digital skill, whereas privacy/risk concerns and preferences for empathy and human interaction were relatively stable across skill levels.

Prior use of AI-based health applications or devices was also associated with more favorable attitudinal profiles. Compared with those reporting no prior use, participants with prior use reported higher exposure, trust, perceived usefulness, and readiness, with small to small–moderate effect sizes (Table 8). Taken together, these patterns suggest that familiarity with AI in health-related contexts and confidence in using digital technologies are linked to greater readiness to accept AI-enabled care, without clear evidence of reduced concern for privacy or diminished emphasis on human interaction. From an implementation perspective, this highlights modifiable levers, including strengthening digital literacy and designing safe, visible applications that build experience and trust, rather than relying primarily on demographic targeting.

Psychometric findings provide additional nuance relevant to measurement and to how acceptability is conceptualized. Trust & transparency and Perceived usefulness showed acceptable internal consistency, supporting their use as distinct constructs in this context. By contrast, Exposure, Privacy & perceived risks, Empathy & human interaction, and Readiness / behavioral intention demonstrated more modest reliability (Table 2), which likely attenuates observed associations and suggests that these domains are more heterogeneous. Item level diagnostics (Table 11) indicated that items explicitly addressing division of labor and authority, such as preferring AI to work alongside rather than replace physicians, concern that AI recommendations could override physician judgment, and preference for joint AI and physician review, were less coherent within their assigned subscales than neighboring items. Conceptually, this pattern is consistent with human oversight or shared control forming a partially distinct dimension of AI acceptability that cuts across empathy, perceived risk, and intention. Future work should refine and validate instruments in Arabic speaking settings to capture this oversight and shared control dimension explicitly and to clarify how it interacts with trust, perceived usefulness, and readiness in shaping patient acceptance of AI enabled care.

**Table 11.** Item diagnostic summary for items reflecting oversight and shared control.

| Item code and English paraphrase | Corrected $r_{it}$ | $\alpha$ if deleted |
| --- | --- | --- |
| PR5: privacy or risk, concern that AI recommendations override physician judgment | 0.24 | 0.685 |
| HI5: empathy or human interaction, prefer AI to work alongside the physician rather than alone | 0.17 | 0.675 |
| BI2: readiness, prefer joint review of my case by AI and the physician | 0.26 | 0.714 |

## Limitations

This study has several limitations. First, the cross sectional design precludes causal inference and all reported associations are correlational. Second, recruitment from selected hospitals in three governorates using non probability sampling limits generalizability to the wider Jordanian population and to other healthcare settings.

Third, all data were based on self report collected through interviewer administered questionnaires, and may be affected by social desirability bias, interviewer effects, and heterogeneous interpretations of AI (for example chatbots, decision support tools, and imaging algorithms). Fourth, several constructs showed only modest internal consistency, and item level diagnostics suggested that oversight and authority related items may represent a partially distinct latent dimension that is not fully captured by the current subscales. Formal psychometric work, including factor analysis and test and retest reliability in Arabic speaking populations, is needed before these scales are used for high stakes comparisons.

Fifth, the regression model explained a small proportion of variance in readiness (*R*^2^ ≈ 0.10) and included only demographic and technology related predictors; other potentially important determinants (for example detailed facets of trust, perceived fairness, prior healthcare experiences, and specific AI use cases) were not assessed. Finally, multiple hypothesis tests were conducted without correction for multiplicity, increasing the risk that some statistically significant findings with small effect sizes reflect type I error (false positive results).

## Conclusion

In this hospital-based sample from Jordan, patients reported relatively high perceived usefulness of AI in healthcare, moderate-to-high trust and readiness to accept AI-enabled care, and strong preferences for empathic, human-centered interaction and clinician involvement. Privacy and perceived risk concerns were present at moderate levels, and self-reported exposure to AI in clinical care was limited. Educational attainment and self-rated digital skill were associated with readiness, suggesting that structural and digital capability factors may shape patient acceptance.

For health system modernization in Jordan and similar settings, these findings support implementing AI primarily as clinician-supportive technology, coupled with clear communication about its use, strong privacy and data-governance safeguards, and targeted digital capability–building efforts for less-educated and lower-skill groups. Future research should refine and validate measures of AI acceptability, explicitly assess oversight and shared-control preferences, and examine how evolving experience, policy, and technology influence patient attitudes over time.

## Data Availability

The data underlying this study contain de-identified human participant survey responses and cannot be made publicly available due to ethical and privacy considerations under the approved study protocol. A minimal, de-identified dataset sufficient to replicate the findings is available from the relevant institutional ethics committee upon reasonable request, subject to approval and in accordance with applicable regulations.

## References

1. Topol EJ. High-performance medicine: the convergence of human and artificial intelligence. Nature Medicine. 2019;25(1):44–56. doi:10.1038/s41591-018-0300-7.

2. World Health Organization. Ethics and governance of artificial intelligence for health: WHO guidance. Geneva: World Health Organization; 2021.

3. World Health Organization. Ethics and governance of artificial intelligence for health: Guidance on large multi-modal models. Geneva: World Health Organization; 2025.

4. U S Food and Drug Administration. Marketing Submission Recommendations for a Predetermined Change Control Plan for Artificial Intelligence-Enabled Device Software Functions. U.S. Food and Drug Administration; 2025. Guidance for industry and FDA staff.

5. Vasey B, Nagendran M, Campbell B, Clifton DA, Collins GS, Denaxas S, et al. Reporting guideline for the early-stage clinical evaluation of decision support systems driven by artificial intelligence: DECIDE-AI. Nature Medicine. 2022;28(5):924–33. doi:10.1038/s41591-022-01772-9.

6. Collins GS, Moons KGM, Dhiman P, Riley RD, Beam AL, Van Calster B, et al. TRIPOD+AI statement: updated guidance for reporting clinical prediction models that use regression or machine learning methods. BMJ. 2024;385:e078378. doi:10.1136/bmj-2023-078378.

7. Sounderajah V, Guni A, Liu X, Collins GS, Karthikesalingam A, Markar SR, et al. The STARD-AI reporting guideline for diagnostic accuracy studies using artificial intelligence. Nature Medicine. 2025;31(10):3283–9. doi:10.1038/s41591-025-03953-8.

8. Gallifant J, Afshar M, Ameen S, Aphinyanaphongs Y, Chen S, Cacciamani G, et al. The TRIPOD-LLM reporting guideline for studies using large language models. Nature Medicine. 2025;31(1):60–9. doi:10.1038/s41591-024-03425-5.

9. Lee JD, See KA. Trust in automation: designing for appropriate reliance. Human Factors. 2004;46(1):50–80. doi:10.1518/hfes.46.1.5030392.

10. Young AT, Amara D, Bhattacharya A, Wei ML. Patient and general public attitudes towards clinical artificial intelligence: a mixed methods systematic review. The Lancet Digital Health. 2021;3(9):e599–611. doi:10.1016/S2589-7500(21)00132-1.

11. Obermeyer Z, Powers B, Vogeli C, Mullainathan S. Dissecting racial bias in an algorithm used to manage the health of populations. Science. 2019;366(6464):447–53. doi:10.1126/science.aax2342.

12. Young AT, Amara D, Bhattacharya A, Wei ML. Patient and general public attitudes towards clinical artificial intelligence: a mixed methods systematic review. The Lancet Digital Health. 2021;3(9):e599–611. doi:10.1016/S2589-7500(21)00132-1.

13. Beets B, Newman TP, Howell EL, Bao L, Yang S. Surveying Public Perceptions of Artificial Intelligence in Health Care in the United States: Systematic Review. Journal of Medical Internet Research. 2023;25:e40337. doi:10.2196/40337.

14. Busch F, Hoffmann L, Xu L, Zhang LJ, Hu B, García-Juárez I, et al. Multinational Attitudes Toward AI in Health Care and Diagnostics Among Hospital Patients. JAMA Network Open. 2025;8(6):e2514452. doi:10.1001/jamanetworkopen.2025.14452.

15. Robertson C, Woods A, Bergstrand K, Findley J, Balser C, Slepian MJ. Diverse patients’ attitudes towards Artificial Intelligence (AI) in diagnosis. PLOS Digital Health. 2023;2(5):e0000237. doi:10.1371/journal.pdig.0000237.

16. Nong P, Ji M. Expectations of healthcare AI and the role of trust: understanding patient views on how AI will impact cost, access, and patient-provider relationships. Journal of the American Medical Informatics Association. 2025;32(5):795–9. doi:10.1093/jamia/ocaf031.

17. Fritsch SJ, Blankenheim A, Wahl A, Hetfeld P, Maassen O, Deffge S, et al. Attitudes and perception of artificial intelligence in healthcare: A cross-sectional survey among patients. Digital Health. 2022;8:20552076221116772. doi:10.1177/20552076221116772.

18. Babitsch B, Hannemann N, Hubner U. Trust in Artificial Intelligence in Wound Care: Perspectives of Healthcare Professionals and Patients in Germany. Studies in Health Technology and Informatics. 2025;332:144–8. doi:10.3233/SHTI251514.

19. Warrington DJ, Holm S. Healthcare ethics and artificial intelligence: a UK doctor survey. BMJ Open. 2024;14(12):e089090. doi:10.1136/bmjopen-2024-089090.

20. Rojahn J, Palu A, Skiena S, Jones JJ. American public opinion on artificial intelligence in healthcare. PLOS ONE. 2023;18(11):e0294028. doi:10.1371/journal.pone.0294028.

21. Temple S, Rowbottom C, Simpson J. Patient views on the implementation of artificial intelligence in radiotherapy. Radiography. 2023;29:S112–6. doi:10.1016/j.radi.2023.03.006.

22. Allam RM, Abdelfatah D, Khalil MIM, Elsaieed MM, El Desouky ED. Medical students and house officers’ perception, attitude and potential barriers towards artificial intelligence in Egypt, cross sectional survey. BMC Medical Education. 2024;24(1):1244. doi:10.1186/s12909-024-06201-8.

23. Amiri H, Peiravi S, Rezazadeh Shojaee SS, Rouhparvarzamin M, Nateghi MN, Etemadi MH, et al. Medical, dental, and nursing students’ attitudes and knowledge towards artificial intelligence: a systematic review and meta-analysis. BMC Medical Education. 2024;24(1):412. doi:10.1186/s12909-024-05406-1.

24. Chen Y, Wu Z, Wang P, Xie L, Yan M, Jiang M, et al. Radiology Residents’ Perceptions of Artificial Intelligence: Nationwide Cross-Sectional Survey Study. Journal of Medical Internet Research. 2023;25:e48249. doi:10.2196/48249.

25. Labrague LJ, Aguilar-Rosales R, Yboa BC, Sabio JB, de Los Santos JA. Student nurses’ attitudes, perceived utilization, and intention to adopt artificial intelligence (AI) technology in nursing practice: A cross-sectional study. Nurse Education in Practice. 2023;73:103815. doi:10.1016/j.nepr.2023.103815.

26. Buabbas AJ, Miskin B, Alnaqi AA, Ayed AK, Shehab AA, Syed-Abdul S, et al. Investigating Students’ Perceptions towards Artificial Intelligence in Medical Education. Healthcare. 2023;11(9):1298. doi:10.3390/healthcare11091298.

27. Allam AH, Eltewacy NK, Alabdallat YJ, Owais TA, Salman S, Ebada MA, et al. Knowledge, attitude, and perception of Arab medical students towards artificial intelligence in medicine and radiology: A multi-national cross-sectional study. European Radiology. 2024;34(7):4494–506. doi:10.1007/s00330-023-10516-z.

28. Daher OA, Dabbousi AA, Chamroukh R, Saab AY, Al Ayoubi AR, Salameh P. Artificial Intelligence: Knowledge and Attitude Among Lebanese Medical Students. Cureus. 2024;16(1):e51876. doi:10.7759/cureus.51876.

29. Baghdadi LR, Mobeirek AA, Alhudaithi DR, Albenmousa FA, Alhadlaq LS, Alaql MS, et al. Patients’ Attitudes Toward the Use of Artificial Intelligence as a Diagnostic Tool in Radiology in Saudi Arabia: Cross-Sectional Study. JMIR Human Factors. 2024;11:e53108. doi:10.2196/53108.

30. Syed W, Babelghaith SD, Al-Arifi MN. Assessment of Saudi Public Perceptions and Opinions towards Artificial Intelligence in Health Care. Medicina. 2024;60(6):938. doi:10.3390/medicina60060938.

31. Jarab AS, Al-Qerem W, Al-Hajjeh DM, Abu Heshmeh S, Mukattash TL, Naser AY, et al. Artificial intelligence utilization in the healthcare setting: perceptions of the public in the UAE. International Journal of Environmental Health Research. 2025;35(3):585–93. doi:10.1080/09603123.2024.2363472.

32. Dalky A, Malkawi RMO, Alrawashdeh A, ALBashtawy M, Bani Hani S. Perceptions, barriers, and risk of artificial intelligence Among healthcare professionals: A cross-sectional study. Digital Health. 2025;11:20552076251360924. doi:10.1177/20552076251360924.

33. Oweidat IA, Alkhatib M, ALBashtawy M, Al Omar S, Al-Rjoub S, Alsaqer K, et al. Knowledge, attitudes, practices, and barriers of artificial intelligence as predictors of intent to stay among nurses: A cross-sectional study. Digital Health. 2025;11:20552076251336106. doi:10.1177/20552076251336106.

34. Al-Qerem W, et al. Artificial intelligence in healthcare education: A cross-sectional study of students’ knowledge, attitudes, and practices. BMC Medical Informatics and Decision Making. 2023;23:288. doi:10.1186/s12911-023-02403-0.

35. Mosleh SM, et al. Knowledge, attitudes, and practices toward artificial intelligence among medicine and pharmacy students in Jordan and the West Bank of Palestine. Advances in Medical Education and Practice. 2023;14:1391–400. doi:10.2147/AMEP.S433255.

36. Hamad S, et al. Medical students’ readiness for artificial intelligence: evidence using the MAIRS-MS readiness scale. Journal of Medical Education and Curricular Development. 2024;11:23821205241281648. doi:10.1177/23821205241281648.

37. Al Saad MM, et al. Medical students’ knowledge and attitudes toward artificial intelligence in Jordan. Open Public Health Journal. 2022.

38. Abufadda M, et al. Radiologists’ and residents’ perspectives on artificial intelligence in Jordan. Informatics in Medicine Unlocked. 2024;46:101538. doi:10.1016/j.imu.2024.101538.

39. Bsisu I, et al. Attitudes of Jordanian anesthesiologists and anesthesia residents towards artificial intelligence: A cross-sectional study. Journal of Personalized Medicine. 2024;14(5):447. doi:10.3390/jpm14050447.

